# Molecular Characterization of Pediatric B-cell Acute Lymphoblastic Leukemia using Integrative Transcriptomics: A Multicenter Study in Argentina

**DOI:** 10.1101/2024.09.19.24313988

**Authors:** María Sol Ruiz, Ezequiel Sosa, Daniel Avendaño, Ignacio Gomez Mercado, María Laura Lacreu, María Cecilia Riccheri, Virginia Schuttenberg, Luis Aversa, Elba Vazquez, Geraldine Gueron, Javier Cotignola

## Abstract

Acute lymphoblastic leukemia (ALL) is the most common childhood cancer worldwide, and exhibits high molecular heterogeneity. Molecular subtypes are characterized by specific chromosomal and molecular alterations, which are critical for guiding risk-adapted therapies. However, the increasing number of recognized prognostic molecular subtypes demands large resources, which are often limited in low/middle-income countries; thereby restricting the molecular characterization. This study aimed to perform an integrated molecular characterization of childhood B-ALL in Argentine patients. We performed RNA-seq on diagnostic bone marrow aspirates from Argentine patients enrolled in the ALLIC-GATLA-2010 protocol. We used different bioinformatic tools to identify and validate single nucleotide variants, fusion transcripts, gene expression profiles and molecular subtypes. We successfully determined transcriptome-based molecular subtype in 93.7% of patients; with high concordance to conventional karyotyping and RT-PCR (17/18 patients with available molecular data). Analysis of chimeric transcripts revealed 82 fusions, both intra– and inter-chromosomal, suggesting that leukemic cells may undergo chromosomal instability. Two of these fusions were novel: *SCAF8::FER1L4* and *DBF4B::EFTUD2*. We also identified 21 different SNVs/InDels in 16 genes, including three novel variants (DUX4 p.I65N, CREBBP p.G1542V, and CSF3R p.G147R) and predicted to alter protein function. Overall, we observed that all patients who relapsed carried high-risk genetic alterations at diagnosis. Whole-transcriptome analysis of leukemic bone marrow enabled molecular subtyping and the identification of both known and novel molecular alterations associated with prognosis.

## Introduction

Acute lymphoblastic leukemia (ALL) is the most common childhood cancer worldwide. In Argentina, ALL accounted for 29.3% of all pediatric cancers diagnosed between 2000 and 2019, with an average of 395 new cases annually and an age-standardized incidence rate of 4.93 per 100,000 individuals^1^.

Advances in chemotherapy regimens and the stratification of patients into risk groups have led to steady improvements in overall and relapse-free survival. However, despite these advancements, 15-30% of patients still relapse, often facing severe treatment complications and poor overall survival rates^2^. Further intensification of traditional chemotherapy is unlikely to yield better outcomes due to the associated acute toxicity and an unfavorable toxicity/benefit ratio^3,4^. Therefore, understanding the intrinsic and extrinsic factors that contribute to disease development and treatment failure is crucial.

Childhood ALL outcomes vary significantly worldwide, with survival rates exceeding 80% in most high-income countries, but dropping to 20% in low– and middle-income countries^5,6,7^. According to the Argentinian Oncopediatric Hospital Registry (ROHA: Registro Oncopediátrico Hospitalario Argentino), three-year survival rates plateaued at approximately 76% (69.0%, 76.0%, and 76.1% for the periods 2000–2005, 2006–2011, and 2012–2016, respectively), with a five-year survival rate of 72.2% between 2005 and 2014^1^.

Poorer outcomes in low– and middle-income countries are generally attributed to limited access to healthcare facilities, lack of standard-of-care diagnostics, and restricted access to effective treatments. Additionally, recent evidence suggests that genetic ancestry may influence the outcome of the molecular subtypes and patient prognosis^8^. However, how regional genetic and environmental factors affect treatment response and disease development remains poorly understood.

B-cell ALL (B-ALL) is the most frequent type of ALL, with 27 molecular subtypes (including 4 provisional entities) recognized by the International Consensus Classification^9^. These molecular subtypes are characterized by specific chromosomal alterations, gene expression profiles, aneuploidies and point mutations^10^, which are crucial for guiding risk-adapted therapies and precision medicine. However, the ability to identify these subtypes accurately depends on the availability of molecular diagnostic techniques, which vary across medical centers. The increasing complexity of molecular classification presents significant technical, analytical, and economic challenges, particularly in low– and middle-income countries, limiting the widespread implementation of these advanced diagnostic approaches^11^.

The aim of this study was to perform an integrated molecular characterization of childhood B-ALL in Argentine patients enrolled in the ALLIC-GATLA-2010 protocol. Using whole-transcriptome sequencing (RNA-seq) on bone marrow aspirates and free user-friendly bioinformatic tools, we aimed to identify known and novel chromosomal rearrangements, small insertions/deletions (InDels) and single nucleotide variants (SNVs), and evaluate their association with clinical outcomes. Additionally, we conducted *in silico* analyses to further explore the novel genetic variants identified. Our findings provide valuable insights into the molecular heterogeneity of B-ALL and establish a robust framework for incorporating transcriptome sequencing into clinical diagnostics.

## Materials and Methods

### Patients and samples

Bone marrow aspirates (1-5 mL) were obtained from newly diagnosed, untreated children and adolescents with ALL enrolled in the Argentine multicenter clinical protocol ALLIC-GATLA 2010. Samples were collected across three hospitals in Argentina: Hospital Nacional Posadas (n=8), Hospital de Niños Dr. Ricardo Gutierrez (n=5), and Hospital Sor María Ludovica (n=19). The clinico-pathological characteristics and disease outcome were evaluated by trained oncohematologists. All procedures involving human participants complied with the ethical guidelines outlined by the Institutional Review Board (IRB) and with the Declaration of Helsinki. The study protocol was also approved by Argentina’s National Drug, Food and Technology Administration (ANMAT).

The inclusion criteria were: 1) IRB protocol approval, 2) *de novo* primary ALL, 3) patient age between 1 and 18 years old, 4) written informed consent from parents or legal guardians, with written informed assent from patients when applicable. Risk stratification was conducted in accordance with the protocol guidelines based on age, white blood cell count at diagnosis, percentage of blasts at day 8, minimal residual disease (MRD) at days 15 and 33 (evaluated by multiparametric flow cytometry), ploidy, t(9;22) *BCR::ABL1*, and t(4;11) *KMT2A::AFF1*.

### Transcriptome sequencing

Frozen cell pellets in Trizol or purified total RNA were shipped to the Inflammation and Cancer Laboratory at Facultad de Ciencias Exactas y Naturales, Universidad de Buenos Aires (CABA, Argentina). Total RNA was isolated using either Trizol (Thermo Fisher Scientific, USA) or Quick-Zol (Kallium technologies, Argentina) following the manufacturer’s protocols, either at the extraction site or in our laboratory. The RNA was resuspended in RNAse/DNase-free distilled water and stored at –80°C. RNA quantification and integrity were evaluated using a Nanodrop (Thermo Fisher Scientific, USA) and Bioanalyzer (Agilent Technologies, USA).

Transcriptome sequencing was performed on 32 bone marrow aspirates collected at the time of diagnosis. RNA-seq libraries were prepared following ribosomal RNA depletion using the Ribo-Zero Plus rRNA Depletion Kit (Illumina, USA). Paired-end RNA sequencing was conducted at Macrogen (Korea) on HiSeq2000 or HiSeq2500 Systems (Illumina, USA) in five separate batches over three years.

### Bioinformatic methods

RNA-seq quality was assessed using *FastQC* (v0.11.5, RRID:SCR_014583). Reads were pseudoaligned to the GRCh38 reference transcriptome using Kallisto (RRID:SCR_016582)^12^ to obtain transcript counts. Transcripts were annotated using the EnsDb.Hsapiens.v86 R package (Bioconductor). SNVs and small InDels were evaluated using RNAmut^13^ based on a custom index of 114 genes and 79 fusion genes associated with ALL (Supplementary Material S1). Variants were annotated and filtered using an in-house pipeline (Supplementary Material S2) with the following criteria: A) number of reads: MutReads >= 15; B) Variant Allele Fraction in RNA: VAF >= 0.2; C) frequency in population databases: Freq_1000genomes < 1% or not available; D) clinical significance in ClinVar (RRID:SCR_006169); E) association with leukemia phenotype; and F) pathogenicity predictions from Polyphen (RRID:SCR_013189). Variants identified in the RNA-seq were confirmed by RT-PCR and Sanger sequencing.

Novel fusion genes were identified using STAR-Fusion (v1.11.0, RRID:SCR_025853)^14^ and FusionInspector^15^. Both tools were run on Singularity at the CCAD-UNC (Centro de Cómputo de Alto Desempeño, Universidad Nacional de Córdoba). Molecular subtypes were predicted with the ALLsorts^16^ and ALLCatchR^17^ classifiers on the high-performance cluster (CeCAR) at Facultad de Ciencias Exactas y Naturales, Universidad de Buenos Aires. Protein structural analysis was performed using the VMD software (RRID:SCR_001820, VMD is developed with NIH support by the Theoretical and Computational Biophysics group at the Beckman Institute, University of Illinois at Urbana-Champaign).

### Statistical analysis and graphics

Statistical analyses, tables and graphics were generated using Rstudio with the following packages: *tidyverse*, *gtsummary*, *plotly, survival, survminer, reshape2* and *chimeraviz*. The oncoprint chart was created using the script available in: https://github.com/sarahet/The_Distinct_DNA_Methylome_ALL/blob/main/Figure5_Extended_Data_Figure5.R.

## Results

### Cohort demographics

The Intercontinental Berlin-Frankfurt-Münster Study Group (IC-BFM-SG) is a cooperative organization that brings together countries from Europe, Latin America and, initially, Asia. The IC-BFM-SG has conducted two large clinical trials in childhood ALL, with a focus on providing support and guidance to countries with limited resources^18^.

The patients included in our study were enrolled under the ALLIC BFM-2009 protocol through the Argentine multicentric protocol ALLIC-GATLA (Grupo Argentino de Tratamiento de la Leucemia Aguda) 2010-2020^19^. We recruited 32 patients diagnosed with primary *de novo* B-ALL. Demographic and clinico-pathological data are shown in Table 1. In summary, the median age at diagnosis was 7 years (range: 2-18 years), with a sex distribution of 47% boys and 53% girls. The studied cohort was composed of 3 patients categorized as standard-risk (9%), 22 as intermediate-risk (69%) and 7 as high-risk (22%). The median follow-up time was 32 months. Overall Survival (OS) and Event-Free Survival (EFS) at 36 months were 84.2% (Standard Error (SE) = 6.5%) and 75.6% (SE = 8.3%), respectively (Figure 1).

**FIGURE 1.**
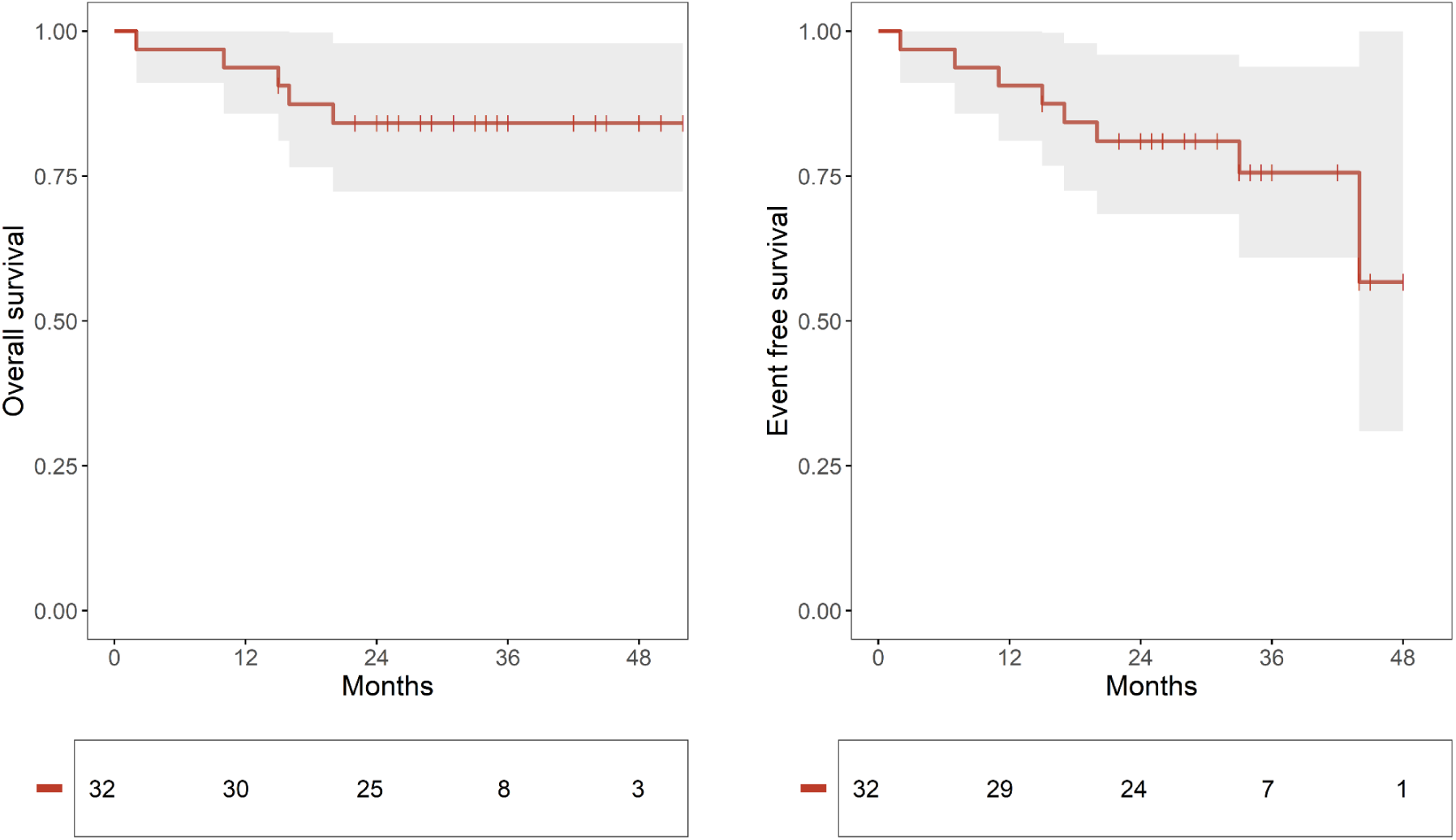
Overall and Event-free survival analyses. The figure displays Kaplan-Meier survival curves for overall survival (left) and event-free survival (right). For event-free survival, the time-to-event was defined as the interval from diagnosis to either relapse or death, whichever occurred first.

**TABLE 1.** Demographic and clinico-pathological data.

| Variable | Total n = 32 |
| --- | --- |
| **Age at diagnosis** |  |
| median (range) | 7.0 (2-18) years |
| IQR=interquartile range | 3-13 years |
| **Sex** |  |
| Female | 17 (53%) |
| Male | 15 (47%) |
| **ALL subtype** |  |
| common | 26 (81%) |
| pre-B | 6 (19%) |
| **Risk group (ALLIC-BFM-2009<sup>19</sup>)** |  |
| standard | 3 (9.4%) |
| intermediate | 22 (68.7%) |
| high | 7 (21.9%) |
| **Response to Prednisone** |  |
| good | 28 (88%) |
| poor | 4 (13%) |
| **Minimal Residual Disease at day 15** |  |
| <0,1% | 11 (37%) |
| 1-10% | 14 (47%) |
| >10% | 5 (17%) |
| Unknown | 2 |
| **Acute severe toxicity** |  |
| Yes | 9 (29%) |
| Unknown | 1 |
| **Relapse** |  |
| Yes | 5 (16%) |
| **Death** |  |
| Yes | 5 (16%) |
| **Follow-up time (months)** |  |
| median (range) | 32 (2-52) months |
| IQR=interquartile range | 25-35 months |

### Molecular subtyping using RNA-seq-based classifiers

We used the ALLSorts classifier, a machine learning algorithm designed to categorize B-ALL patients into 18 predefined molecular subtypes based on whole transcriptome sequencing data^16^. Since ALLSorts indicates that misclassification often occurs with the “high hyperdiploid”, “low hyperdiploid”, and “near haploid” subtypes, and classification accuracy improves when using the meta-subtype “High Sig”^16^, we opted to use this meta-subtype and called it “hyperdiploid”. ALLSorts successfully assigned the molecular subtype to 30 out of the 32 patients (93.7%); the remaining 2 were categorized as B-other/unclassified (Figure 2A and Table 2). These results showed high concordance with conventional subtyping methods like karyotyping and RT-PCR (with 17/18 samples (94%) with available cytogenetic and molecular tests). Of note, the 2010-2020 guidelines for childhood ALL subtyping required testing for only the five most frequent and validated prognostic subgroups (*ETV6::RUNX1*, *BCR::ABL1* (p190 and p210), *KMT2A::AFF1*, hyperdiploid and hypodiploid), limiting the confirmation of the 13 additional subtypes assigned by ALLSorts. We observed one discordant result, a sample that was classified as a DUX4 by ALLSorts and as hyperdiploid by traditional karyotyping. Because chromosome 4, where *DUX4* is located, is frequently affected in the hyperdiploid subtype^20^, this might have contributed to the classification as hyperdiploid by karyotyping and as DUX4 by whole transcriptome analysis. Therefore, we classified this patient as a DUX4/Hyperdiploid molecular subtype and considered it as a high-risk leukemia as per protocol guidelines (MRD>10% at day 15). In summary, we identified 9 B-ALL subtypes in our cohort, with hyperdiploid (37.5%), DUX4 (12.5%) and ETV6::RUNX1 (9.4%) being the most frequent (Figure 2 and Table 2). In addition, the use of RNA-seq followed by ALLSorts increased the molecular subtyping rate from 31.2% (limited by the mandatory molecular and genetic tests in the ALLIC-GATLA-2010 protocol) to 93.7% (Table 2). These frequencies are comparable to those reported by Campbell *et al.* for the ALLIC-BFM-2009 cohort and by Brady *et al.* for the St. Jude cohort^10^ (Table 2).

**FIGURE 2.**
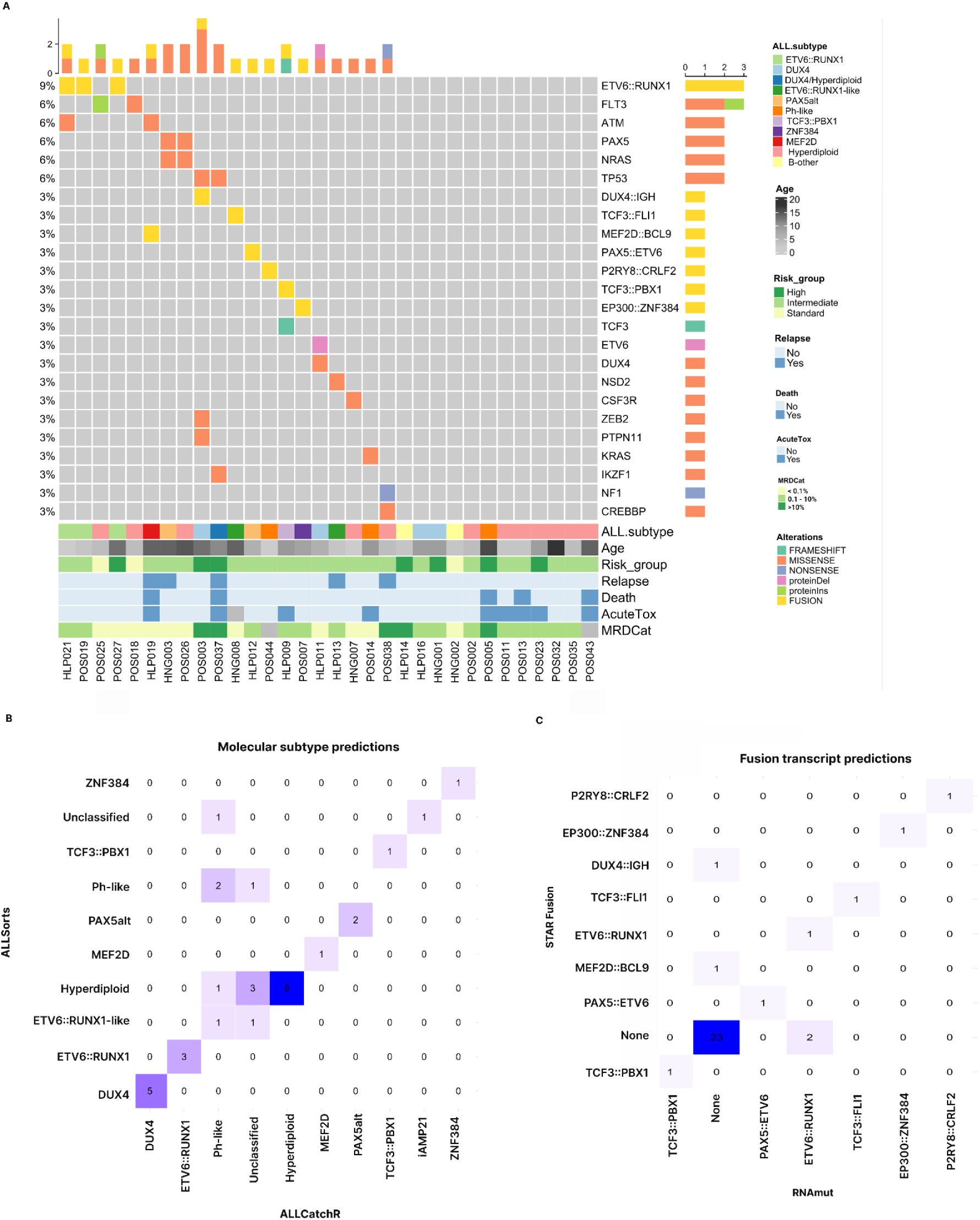
Summary of molecular alterations identified in this study. (A) Overview of the genomic landscape across the cohort, where each column represents an individual patient and each row corresponds to a specific genetic alteration. SNVs, InDels, and fusion transcripts are displayed, ordered by their frequency within the cohort. Molecular subtypes and clinical features (MRDCat: minimal residual disease at day 15) are shown below the chart. (B) Confusion matrix comparing molecular subtyping results from ALLSorts and ALLCatchR. (C) Confusion matrix comparing the identification of fusion transcripts by STAR-Fusion and RNAmut. Only fusion transcripts previously reported in ALL were included in these analyses.

**TABLE 2.**
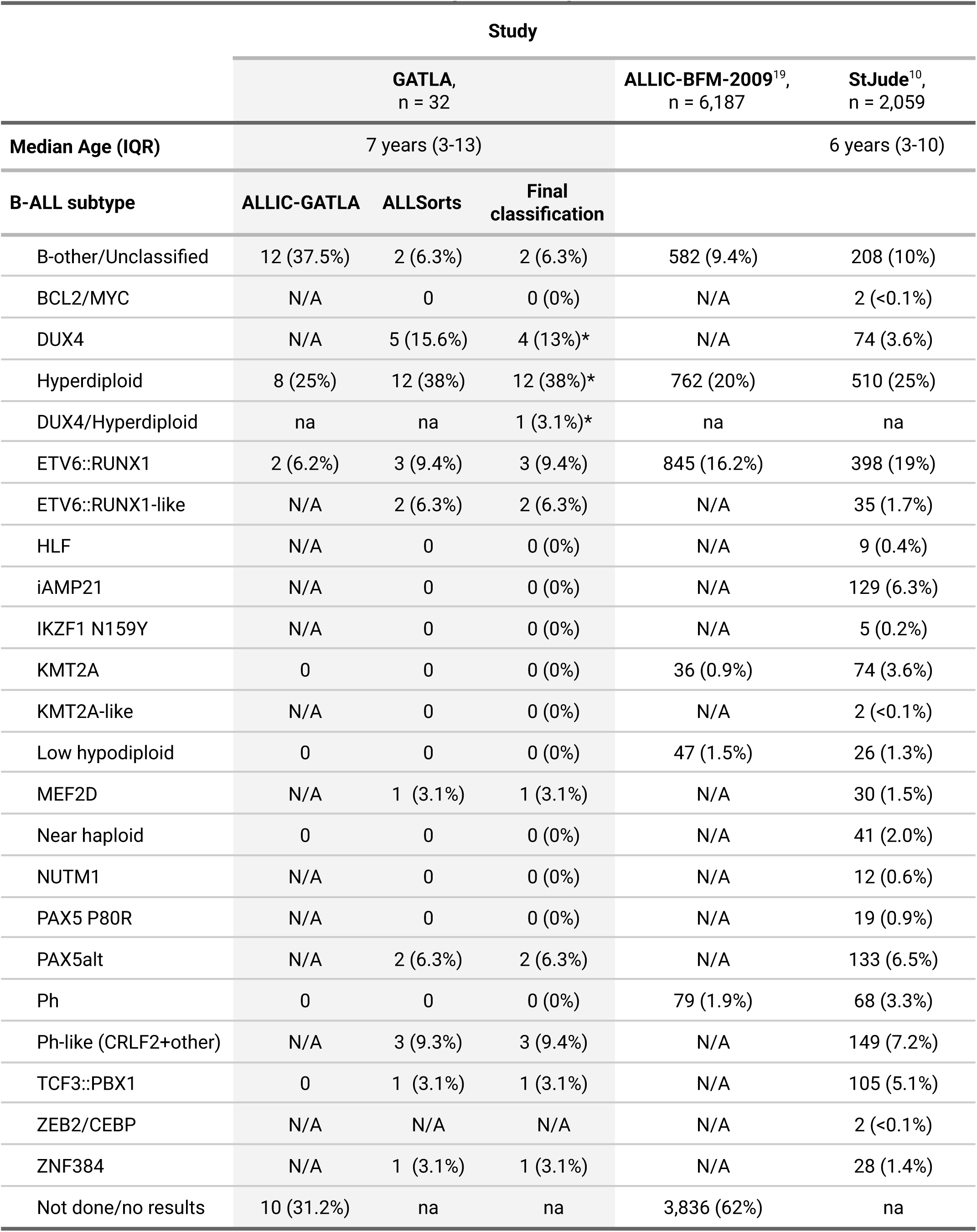
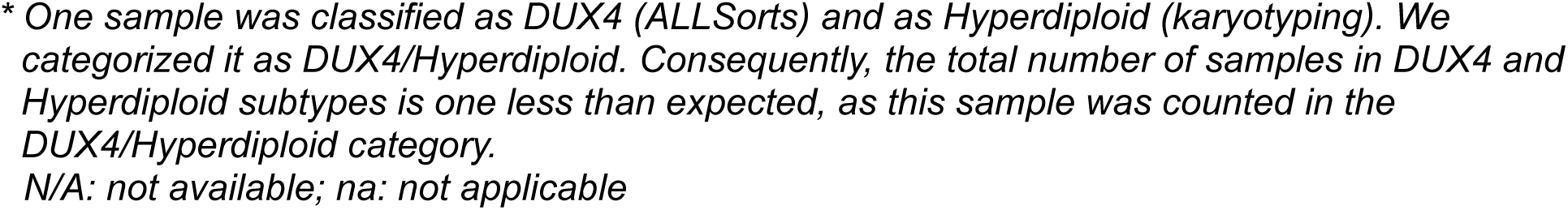
Table summarizing molecular subtypes in this study and their comparison with ALLIC-BFM-2009 and St. Jude-reported frequencies.

Given the difficulty in evaluating the accuracy of subtype predictions in the samples that remained unclassified by current traditional molecular and cytogenetic diagnosis, we used an additional subtype classifier, ALLCatchR^17^. Overall, we found an 82.1% agreement with ALLsorts (23 out of 28 classified samples) (Figure 2B); and 100% agreement for the 19 “high confidence” predictions (Supplementary Material S1). ALLCatchR was unable to classify five samples (15.6%).

### Study of fusion transcripts

We first analyzed the RNA-seq data using STAR-Fusion. While this tool provides high accuracy in fusion transcript detection because it aligns reads to the entire genome^14^, it demands substantial computational resources and processing time, often limiting its use in daily diagnosis settings. STAR-fusion identified a total of 82 chimeric transcripts: 19 (23%) inter-chromosomal, and 63 (77%) intra-chromosomal with an intergenic distance that ranged from <100 kb to >1 Mb (Supplementary Material S1), suggesting intra-chromosomal deletions. Eight of these fusions were previously reported in ALL (*EP300::ZNF384, ETV6::RUNX1, PAX5::ETV6, TCF3::FLI1, TCF3::PBX1, P2RY8::CRLF2, DUX4::IGH, MEF2D::BCL9*) and are associated with prognostic molecular subtypes^21^ (Figure 2A).

We also used RNAmut, a tool designed to detect cancer-specific mutations and fusions from RNA-seq data^13^. Compared to STAR-Fusion, RNAmut requires a pre-build index including the list of genes to be analyzed, resulting in the need of fewer computational resources. These features make it a potentially valuable tool for clinical applications. On the other hand, RNAmut is unable to analyze mutations and fusions in genes not included in the index, non-coding transcripts, transcripts without consensus CDS, and highly variable regions such as *IGH* genes. Therefore, only 24 of the 82 fusions detected by STAR-Fusion were evaluable by RNAmut.

Then, we built a RNAmut index including these 24 RNAmut-evaluable fusions and 77 additional fusions reported in the literature (Supplementary Material S1). Overall, the agreement between RNAmut and STAR-Fusion was low (8/37 in 32 patients, 21.6%), but the concordance increased when we considered only the fusions commonly found in ALL (Figure 2C, Supplementary Material S1). RNAmut did not detect *MEF2D::BCL9*, whereas STAR-Fusion failed to detect *ETV6::RUNX1* in 2 of 3 samples (Figure 2C).

### Identification of chimeras with long non-coding RNA partners, pseudogenes and novel fusion transcripts

After visually reviewing the mapped reads of fusion transcripts identified by STAR-Fusion, we confirmed 13 chimeras involving long non-coding RNAs. Among these, only the *ERG::LINC01423* fusion was highly expressed (65 reads spanning the junction), possibly originated from a 112,000 bp deletion on chromosome 21 that would lead to a partial deletion of *ERG* encompassing exons 2 to 10 (NM_182918.4) (Supplementary Material S2). A similar fusion involving these two genes has been previously reported in the DUX4 subtype in B-ALL patients^22^. In agreement with this, ALLsorts predicted a DUX4 subtype for the patient with the *ERG::LINC01423* fusion.

We also identified one inter-chromosomal fusion involving the *SCAF8* protein-coding gene and the pseudogene *FER1L4* (*SCAF8::FER1L4*; Figure 3A) with 17 reads spanning the breakage points. Because this was a novel fusion, we further analyzed the expression of the two partners. We observed that *FER1L4* was overexpressed (Z-score=5.1), while *SCAF8* expression was not significantly altered (Z-score=0.9) (Figure 3B). We detected *SCAF8::FER1L4* in co-occurrence with the *TCF3::FLI1* fusion –t(11;19)-previously reported as an oncogenic driver in pediatric B-lymphoblastic leukemia/lymphoma^23^. These fusions were found in a 14-year-old patient diagnosed with an intermediate-risk B-ALL according to the ALLIC-GATLA-2010 guidelines, and classified as ETV6::RUNX1-like by ALLSorts.

**FIGURE 3.**
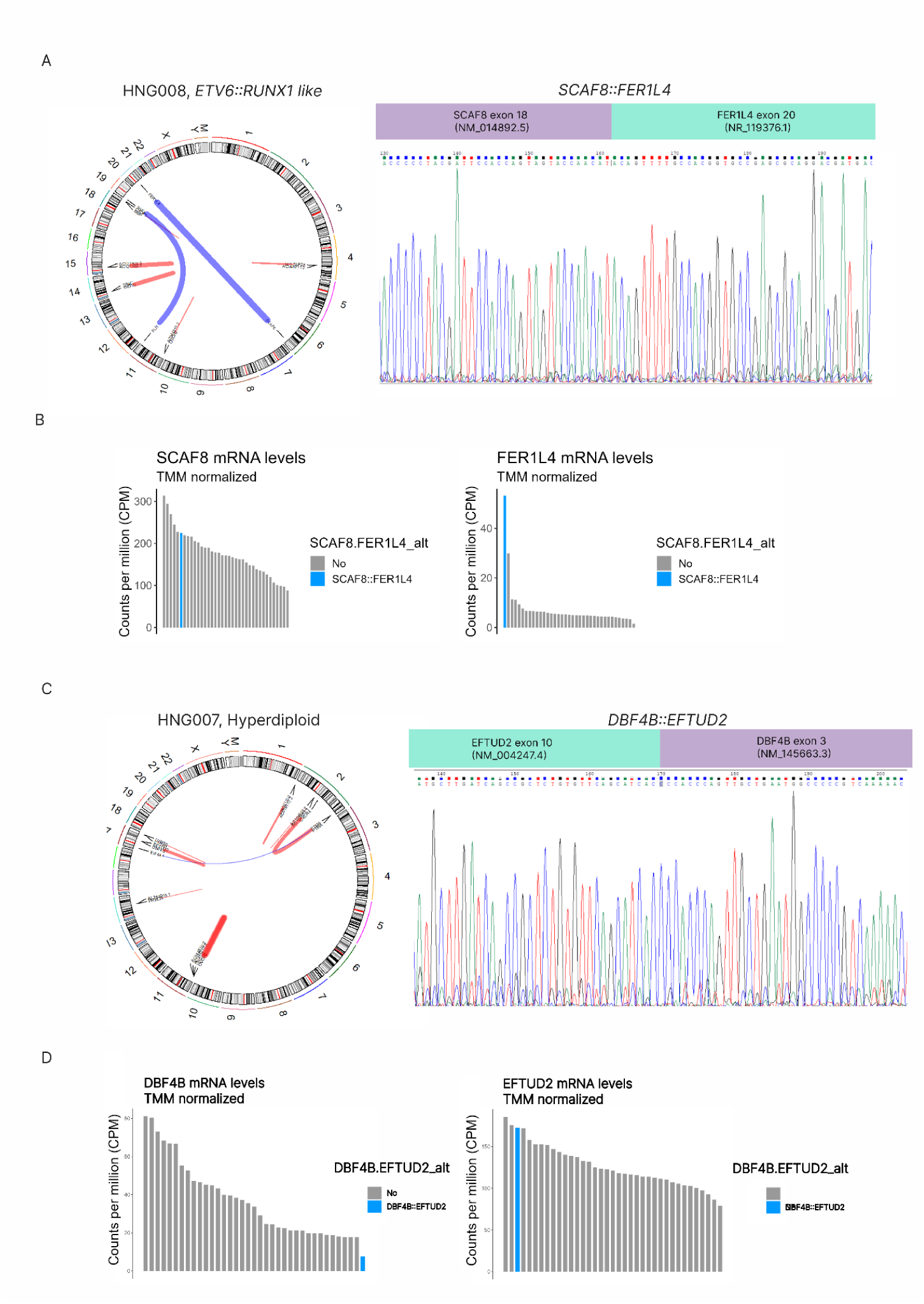
Identification and confirmation of novel fusion transcripts. (A) Circos Plot from a patient classified as ETV6::RUNX1-like. This patient had two translocations (blue lines): t(6;20) *SCAF8::FER1L4* and t(11;19) *TCF3::FLI*; and four intrachromosomal rearrangements (red lines). *SCAF8::FER1L4* novel fusion transcript involved exon 18 of *SCAF8* and exon 20 of *FER1L4*, as confirmed by Sanger sequencing. (B) Analysis of *FER1L4* and *SCAF8* mRNA levels by RNA-seq in the sample harbouring *SCAF8:FER1L4* compared to other patients. This study suggests an overexpression of *FER1L4* levels (Z-score=5.1). (C) Circos Plot from a patient classified as hyperdiploid. This patient had the *DBF4B::EFTUD2* fusion (intrachromosomal, chromosome 17) and multiple additional intrachromosomal rearrangements. This novel fusion transcript involved exon 3 of *DBF4B* and exon 10 of *EFTUD2*, as confirmed by Sanger sequencing. (D) *DBF4B* mRNA levels by RNA-seq suggest a loss of the expression of the *DBF4B*.

Finally, STAR-Fusion and RNAmut detected two novel fusions involving protein-coding genes. The *DBF4B::EFTUD2* fusion was detected with a medium/low sequencing depth (8 reads spanning the junction) and would be the consequence of a ∼110,000 bp intra-chromosomal deletion in chromosome 17. This fusion joins exon 3 of *DBF4B* to exon 10 of *EFTUD2* (Figure 3C) and results in an in-frame chimera. It was identified in a 2-year-old patient with a hyperdiploid subtype and classified as intermediate-risk according to the protocol guidelines. This patient also harbored a SNV in *CSF3R* (p.G147R, described in the following section). The mRNA levels of *DBF4B* suggest a nearly-null expression of wild-type *DBF4B* (Z-score=-1.5) while *EFTUD2* expression was among the highest in this cohort (Z-score=1.8) (Figure 3D).

The other novel fusion, *ABHD17B::CEMIP2*, would be the consequence of a ∼150,000 bp intra-chromosomal deletion in chromosome 9. We detected this fusion in 5 patients, with low abundance (3 to 6 reads), and it was detected by both tools in only one patient. Given the low concordance and expression level, we did not further analyze this fusion.

Finally, we detected chimeric genes in all 32 samples with variable sequencing depth (Supplementary Material S2). Most fusions were intra-chromosomal, likely resulting from chromosomal deletions. While their biological and oncogenic roles remain unknown, this finding suggests that leukemic cells may undergo chromosomal instability during transformation and disease progression. However, we cannot rule out that some of these fusions, particularly those identified at low sequencing depth, may represent noise or spurious transcripts. Alternatively, low-depth fusions could reflect the presence of minor leukemic subclones, highlighting heterogeneity within the leukemic cell population.

### Identification of SNVs and small InDels by transcriptome analysis

We ran RNAmut with a custom index of 114 genes commonly mutated in pediatric ALL to identify SNVs and small InDels (Supplementary Material S1). This analysis detected 9,594 variants across the 32 patients. After annotation and filtering, we identified 21 different SNVs/InDels in 16 genes across 14 patients (42%) (Figure. 2). Based on the OncoKB classification, these SNVs/InDels included 5 oncogenic variants (NSD2 p.E1099K, PTPN11 p.D61F, FLT3 p.N676K, FLT3 p.D835E, and NRAS p.G12D), 7 likely oncogenic variants (ATM p.P604S in two patients, NRAS p.G12S, KRAS p.Q61P, TP53 p.P152L, TP53 p.R267P, NF1 p.R1306*, TCF3 p.S338fs*10), and 9 unclassified variants (PAX5 p.P34L, PAX5 p.R38C, ZEB2 p.H1038R, IKZF1 p.D186Y, CREBBP p.G1542V, DUX4 p.I65N, CSF3R p.G147R, ETV6 p.Q12del, FLT3 R833-D834_Ins:S). Of these, 20 variants were confirmed using CTAT-Mutations, a bioinformatic tool designed for detecting variants from whole-transcriptome data^24^ (Supplementary Material S1); 3 of thems were novel, while the remaining 17 had been previously reported in the literature. The variant detected only by RNAmut (ETV6 p.Q12del) is compatible with an alternative splicing isoform of *ETV6*.

Next, we assessed the potential impact of the 3 novel variants (DUX4 p.I65N, CREBBP p.G1542V, and CSF3R p.G147R) using *in silico* methods. All these variants were located in highly conserved residues and, therefore, were predicted to affect protein function (Table 3, Figure 4, Supplementary Material S2). DUX4 is a transcription factor involved in the regulation of critical genes for early development. The variant p.I65N is located in homeodomain 1, and the residue change likely alters the interaction between DUX4 and its target DNA (Figure 4A), potentially affecting its transcriptional activity.

**FIGURE 4.**
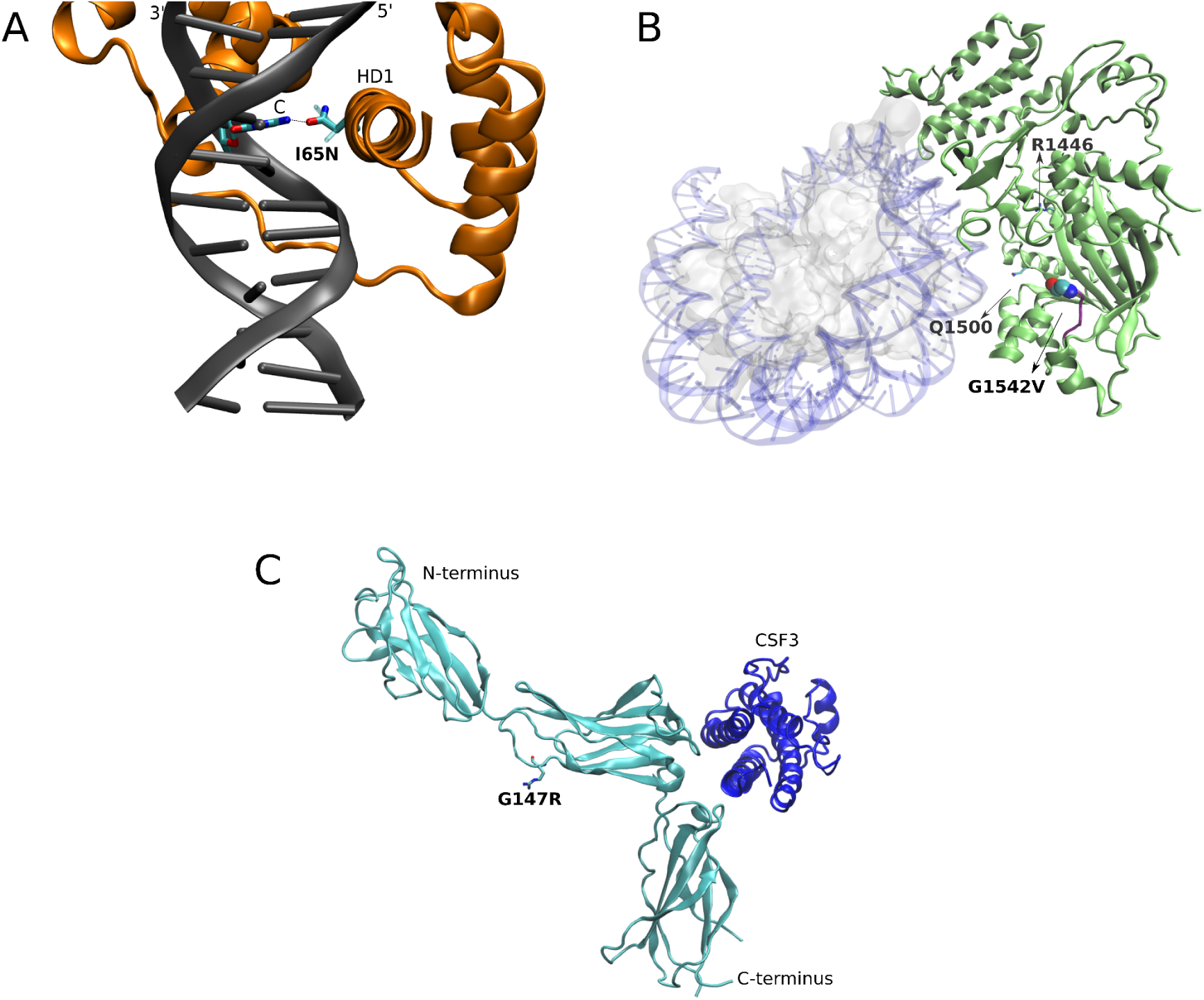
Structural analysis of novel single nucleotide variants. Superimposition of wildtype structure with mutated residues in the described variants are shown. (A) DUX4 a3 helix of HD1 domain inserted in the DNA major groove, with the side chain of residue 65 making base-specific contacts (sticks). Hydrogen bonds are depicted by a dotted line. Note that, while Ile65 in the wild-type (ghost licorice side-chain) DUX4 version does not directly interact with any base, Asn65 variant (solid licorice side-chain) offers new interactions with the base, affecting the original interactions, since the N in the base is now able to form hydrogen bond interactions with the Asn side chain groups, as depicted. PDBid: 5Z6Z. (B) Cryo-EM structure (PDBid: 8HAL) of the CBP catalytic core of CREB-binding protein (green) bound to histone H2B and acetylated H4 (ghost gray) and DNA (transparent blue). As noted, G1542V variant occurs in a disordered region (purple) connecting two ɑ-helices. While not directly located in the protein-histone complex interface, the variant might compromise the stability of this region, as also reported for structurally near variants identified by Mullighan *et al*^25^. (C) CSF3R (cyan) interacting with CSF3 ligand (blue). As noted, while G147R variant (R side chain overlaps with the short G side chain, which is not visible) does not directly affect the intermolecular interaction with CSF3, but it might be responsible for: a destabilization of the loop, an alteration in the interaction with other molecules, or a combination of both. PDBid: 2D9Q

**TABLE 3.** Description of novel single nucleotide variants by pathogenicity/oncogenicity predictors and disease databases.

| Gene | Localization | Variant | Reads var/wt | VAF* | Type of variant | FRANKLIN** | Varsome* | OncoKB | ClinVar | COSMIC | Cancer Genome Interpreter |
| --- | --- | --- | --- | --- | --- | --- | --- | --- | --- | --- | --- |
| *CREBBP* | HAT (histone acetylase protein) domain | NP_004371.2: p.Gly1542Val | 39/0 | 1 | missense | LP | LP | NA | NA | NA, G1542S, G1542D | Driver |
| *CSF3R* | extracellular domain | NP_000751.1: p.Gly147Arg | 92/82 | 0.52 | missense | VUS | VUS | NA | NA | 1 report in basal cell carcinoma | Driver |
| *DUX4* | homeodomain, DNA interaction site | NP_001292997.1: p.Ile65Asn | 657/0 | 1 | missense | VUS | LB | NA | NA | No substitutions in DUX4. | NA |

\*in RNA-seq, \*\*germline classification.
LB: likely benign; LP: likely pathogenic; VAF: variant allele frequency; VUS: variant of uncertain significance; NA: not available.

CREBBP regulates cell growth, division, maturation and differentiation. The novel variant p.G1542V is located in the histone acetyltransferase domain, a known mutation hotspot in relapsed ALL^25^. The p.G1542 residue lies in a disordered region, and the substitution to valine might compromise domain stability and/or interactions with other molecules (Figure 4B).

CSF3R participates in granulopoiesis during inflammation and in cell surface adhesion or recognition processes. The p.G147R variant is located in the receptor’s extracellular region. Although not predicted to directly interfere with the interaction with the ligand CSF3, the substitution of a small uncharged glycine with a larger positive-charged arginine might destabilize the loop, modify the interaction with other molecules, or both (Figure 4C).

### Molecular profiling confirms the accuracy of the bioinformatic subtyping

Whole-transcriptome sequencing provides an integrated view of fusion transcripts, SNVs/InDels and gene expression, allowing for deep molecular subtyping. In order to confirm the accuracy of the bioinformatic subtyping tools used, we analyzed the key biomolecules defining the different ALL subtypes.

The patient classified as ZNF384-alt by ALLSorts harbored the *EP300::ZNF384* fusion transcript and a gene expression profile consistent with the expected immunophenotype: low CD10 (Z-score=-1.14) and high CD33 expression (Z-score=3.3) (Figure 5A).

**FIGURE 5.**
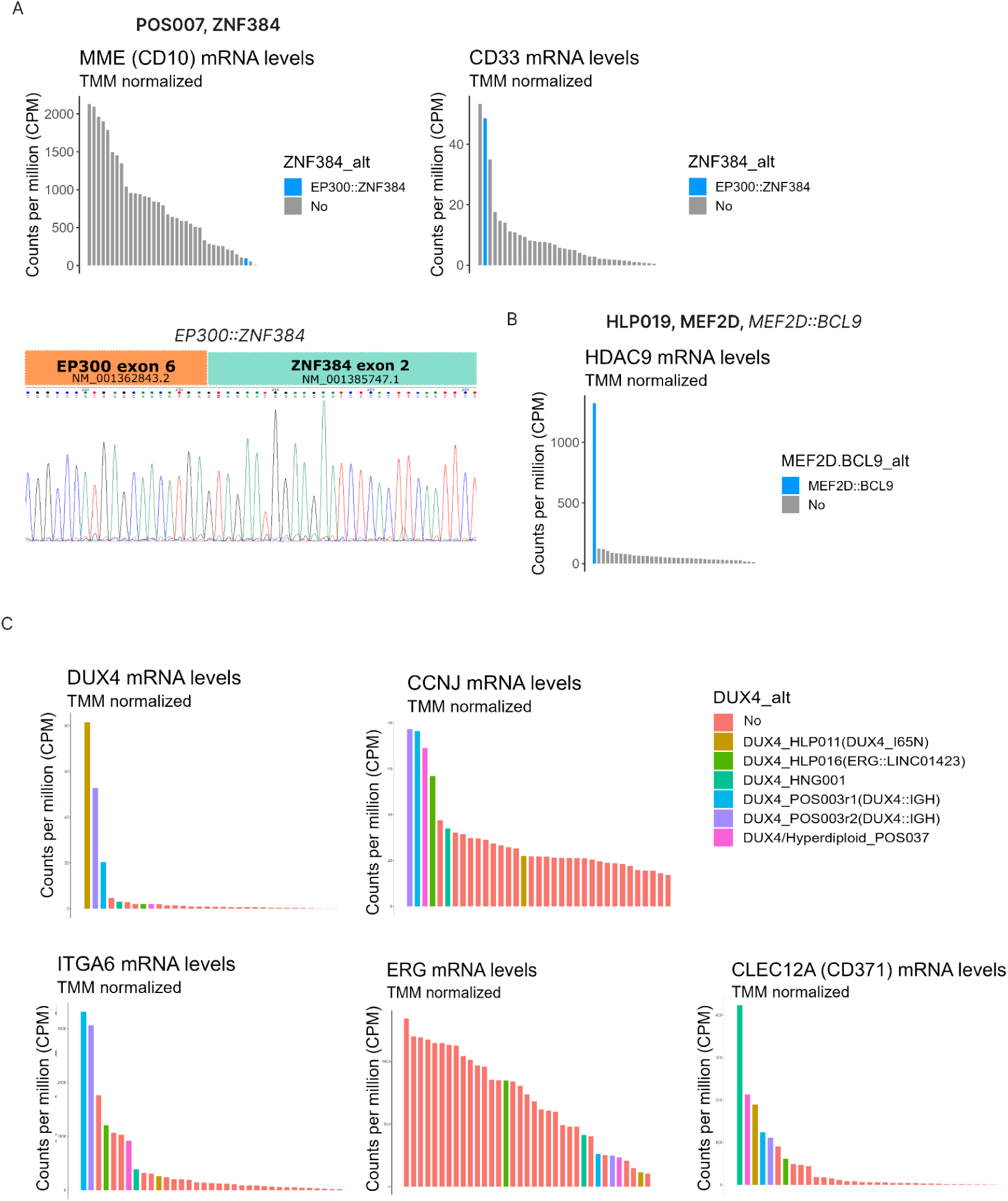
Integration of multiple molecular data confirms the accuracy of the bioinformatic subtyping. ZNF384 subtype was predicted in a patient harboring *EP300::ZNF384* fusion transcript, reduced abundance of *CD10* and increased *CD33* (A). MEF2D was predicted in a patient harboring *MEF2D::BCL9* fusion transcript and increased levels of *HDAC9* mRNA (B). DUX4 subtype was predicted in 5 patients with different molecular features. In some but not all patients there was an association with upregulation of *DUX4::IGH* target genes (*ITGA6*, *CCNJ*), increased levels of *DUX4* and *CD371*, and reduced levels of *ERG* (C).

The patient classified as MEF2D had, as expected, a fusion involving MEF2D (*MEF2D9::BCL9*) and high expression of *HDAC9* (Figure 5B). Although classified as intermediate-risk following the ALLIC-BFM criteria, the patient relapsed at 11 months and died at 16 months, reflecting the poor prognosis associated with this subtype despite an initial good clinical response (MRD<0.1% at days 8 and 15).

ALLSorts identified five patients as DUX4; however, the canonical *DUX4::IGH* fusion transcript was detected only in one sample. As expected, this patient had high expression of *DUX4*, *CCNJ* and *ITGA6*; being the last two direct transcriptional targets of *DUX4::IGH* (Figure 5C). In addition, this patient had mutations in *ZEB2*, *TP53*, and *PTPN11* (Figure 2). Another patient had the novel DUX4 p.I65N variant (Table 3), located within the homeodomain coding region as described in the previous section (Figure 4). This patient showed overexpression of *DUX4*; however, we observed, as predicted by the *in silico* analysis, normal levels of *CCNJ* and *ITGA6* (Figure 5C) suggesting a distinct expression profile compared to the *DUX4::IGH* samples. Another patient carried an intra-chromosomal *ERG* fusion transcript, indicative of a 112,000 bp deletion in chromosome 21 leading to partial *ERG* loss, a common alteration in the DUX4 subtype. Although non-canonical *DUX4* alterations were detected in the remaining cases, all five DUX4 samples had high expression of *CD371* (above quartile 75%), a marker previously associated with this subtype and linked to lineage switching (Figure 5C).

Altogether, these findings confirmed the accuracy of the transcriptome-based computational subtyping tools used in this study.

### Association of molecular alterations with clinical variables

We did not detect significant associations between molecular subtypes and the clinical variables relapse, minimal residual disease (MRD) at day 15, death, response to prednisone at day 8, severe acute toxicity related to treatment, sex nor age. However, given the small sample size in each subtype and the follow-up time (Table 1), we cannot rule out the existence of associations that cannot be detected in this small cohort.

Four of the five patients who relapsed were classified as B-other ALL and one as hyperdiploid following the ALLIC-GALTA-2010 guidelines. Among them, one was diagnosed with high-risk ALL and MRD>10%, one with intermediate-risk disease and MRD>10%, and three with intermediate-risk and MRD<10%. The patient with intermediate-risk disease and MRD>10% harbored a truncating mutation in *NF1* and the novel mutation CREBBP p.G1542V, which we predicted to alter domain stability and/or interactions with other molecules. Notably, all three intermediate-risk patients with MRD<10% had high-risk molecular alterations or subtypes (Tables 4 and 5): NSD2 p.E1099K^26^, *MEF2D::BCL9*^26,27^ / ATM p.P604S (this patient died 16 months after diagnosis), and PAX5alt subtype^28^ (with PAX5 p.P34L) / NRAS p.G12S. Remarkably, two of the latter three patients had co-occurrence of 2 oncogenic mutations. These findings emphasize the importance of comprehensive molecular subtyping to refine risk stratification and guide precision medicine.

**TABLE 4.** Analysis of relapse stratified by molecular subtypes and genetic alterations.

|  |  | Relapse |  | p-value |
| --- | --- | --- | --- | --- |
|  |  | No; n = 27 | Yes; n = 5 |  |
| Follow-up time Median (IQR) |  | 31 (25-35) | 34 (16-50) | 0.70<sup>2</sup> |
| Molecular subtype<sup>1</sup> |  |  |  | 0.24<sup>3</sup> |
|  | Genetic Alteration(s) |  |  |  |
| Hyperdiploid | ABHD17B::CEMIP2<sup>5</sup> | 1 (3.7%) | 0 (0.0%) |  |
|  | CSF3R p.G147R + DBF4B::EFTUD2<sup>5</sup> | 1 (3.7%) | 0 (0.0%) |  |
|  | FLT3 p.N676K | 1 (3.7%) | 0 (0.0%) |  |
|  | FLT3 p.D835E + R833-D834_Ins:S | 1 (3.7%) | 0 (0.0%) |  |
|  | Hyperdiploid | 6 (22.2%) | 0 (0.0%) |  |
|  | NRAS p.G12D + PAX5 p.R38C | 1 (3.7%) | 0 (0.0%) |  |
|  | NF1 p.R1306* + CREBBP p.G1542V | 0 (0.0%) | 1 (20.0%) |  |
|  | DUX4<sup>4</sup> + TP53 p.R267P + IKZF1 p.D186Y | 0 (0.0%) | 1 (20.0%) |  |
|  | total | 11 (40.7%) | 2 (40.0%) |  |
| ETV6::RUNX1 | ETV6::RUNX1 | 2 (7.4%) | 0 (0.0%) |  |
|  | ETV6::RUNX1 + ATM p.P604S | 1 (3.7%) | 0 (0.0%) |  |
|  | total | 3 (11.1%) | 0 (0.0%) |  |
| ETV6::RUNX1-like | NSD2 p.E1099K | 0 (0.0%) | 1 (20.0%) |  |
|  | TCF3::FLI1 + SCAF8::FER1L4<sup>5</sup> | 1 (3.7%) | 0 (0.0%) |  |
|  | total | 1 (3.7%) | 1 (20.0%) |  |
| Ph-like | KRAS p.Q61P | 1 (3.7%) | 0 (0.0%) |  |
|  | Ph-like | 1 (3.7%) | 0 (0.0%) |  |
|  | P2RY8::CRLF2 | 1 (3.7%) | 0 (0.0%) |  |
|  | total | 3 (11.1%) | 0 (0.0%) |  |
| PAX5alt | NRAS p.G12S + PAX5 p.P34L | 0 (0.0%) | 1 (20.0%) |  |
|  | PAX5::ETV6 | 1 (3.7%) | 0 (0.0%) |  |
|  | total | 1 (3.7%) | 1 (20.0%) |  |
| DUX4 | DUX4 | 1 (3.7%) | 0 (0.0%) |  |
|  | DUX4::IGH + PTPN11 p.D61Y + TP53 p.P152L + ZEB2 p.H1038R | 1 (3.7%) | 0 (0.0%) |  |
|  | DUX4 p.I65N + ETV6_Q12del | 1 (3.7%) | 0 (0.0%) |  |
|  | ERG::LINC01423 | 1 (3.7%) | 0 (0.0%) |  |
|  | total | 4 (14.8%) | 0 (0.0%) |  |
| MEF2D | MEF2D::BCL9 + ATM p.P604S | 0 (0.0%) | 1 (20.0%) |  |
| TCF3::PBX1 | TCF3::PBX1 + TCF3_S338fs*10 | 1 (3.7%) | 0 (0.0%) |  |
| ZNF384 | EP300::ZNF384 | 1 (3.7%) | 0 (0.0%) |  |
| B-other | Unclassified | 2 (7.4%) | 0 (0.0%) |  |

<sup>1</sup>Final classification; <sup>2</sup>Wilcoxon rank sum test; <sup>3</sup>Fisher's exact test (by molecular subtype); <sup>4</sup>Classified as DUX4/Hyperdiploid; <sup>5</sup>Novel fusion transcripts. In bold are depicted the high-risk alterations identified in patients initially classified as intermediate-risk according to the protocol guidelines.

**TABLE 5.** Analysis of minimal residual diseases at day 15 by flow cytometry stratified by molecular subtypes and genetic alterations.

|  |  | MRD d15<sup>1</sup> |  |  | p-value |
| --- | --- | --- | --- | --- | --- |
|  |  | <0,1%<br>N = 11 | 1-10%<br>N = 14 | >10%<br>N = 5 |  |
| Follow-up time Median (IQR) |  | 28 (25-33) | 35 (32-36) | 31 (15-48) | 0.20<sup>3</sup> |
| Molecular subtype<sup>2</sup> |  |  |  |  | 0.90<sup>4</sup> |
|  | Genetic Alteration(s) |  |  |  |  |
| Hyperdiploid | ABHD17B::CEMIP2<sup>6</sup> | 0 (0.0%) | 1 (7.1%) | 0 (0.0%) |  |
|  | CSF3R p.G147R + DBF4B::EFTUD2<sup>6</sup> | 1 (9.1%) | 0 (0.0%) | 0 (0.0%) |  |
|  | FLT3 p.N676K | 1 (9.1%) | 0 (0.0%) | 0 (0.0%) |  |
|  | FLT3 p.D835E + R833-D834_Ins:S | 1 (9.1%) | 0 (0.0%) | 0 (0.0%) |  |
|  | Hyperdiploid | 0 (0.0%) | 5 (35.7%) | 0 (0.0%) |  |
|  | NRAS p.G12D + PAX5 p.R38C | 1 (9.1%) | 0 (0.0%) | 0 (0.0%) |  |
|  | NF1 p.R1306* + CREBBP p.G1542V | 0 (0.0%) | 0 (0.0%) | 1 (20.0%) |  |
|  | DUX4<sup>5</sup> + TP53 p.R267P + IKZF1 p.D186Y | 0 (0.0%) | 0 (0.0%) | 1 (20.0%) |  |
|  | total | 4 (36.4%) | 6 (42.9%) | 2 (40.0%) |  |
| ETV6::RUNX1 | ETV6::RUNX1 | 1 (9.1%) | 1 (7.1%) | 0 (0.0%) |  |
|  | ETV6::RUNX1 + ATM p.P604S | 0 (0.0%) | 1 (7.1%) | 0 (0.0%) |  |
|  | total | 1 (9.1%) | 2 (14.3%) | 0 (0.0%) |  |
| ETV6::RUNX1-like | NSD2 p.E1099K | 0 (0.0%) | 1 (7.1%) | 0 (0.0%) |  |
|  | TCF3::FLI1+ SCAF8::FER1L4<sup>6</sup> | 1 (9.1%) | 0 (0.0%) | 0 (0.0%) |  |
|  | total | 1 (9.1%) | 1 (7.1%) | 0 (0.0%) |  |
| Ph-like | KRAS p.Q61P | 1 (9.1%) | 0 (0.0%) | 0 (0.0%) |  |
|  | Ph-like | 0 (0.0%) | 0 (0.0%) | 1 (20.0%) |  |
|  | P2RY8::CRLF2 | NA |  |  |  |
|  | total | 1 (9.1%) | 0 (0.0%) | 1 (20.0%) |  |
| PAX5alt | NRAS p.G12S + PAX5 p.P34L | 1 (9.1%) | 0 (0.0%) | 0 (0.0%) |  |
|  | PAX5::ETV6 | 0 (0.0%) | 1 (7.1%) | 0 (0.0%) |  |
|  | total | 1 (9.1%) | 1 (7.1%) | 0 (0.0%) |  |
| DUX4 | DUX4 | 0 (0.0%) | 1 (7.1%) | 0 (0.0%) |  |
|  | DUX4::IGH + PTPN11 p.D61Y + TP53 p.P152L + ZEB2 p.H1038R | 0 (0.0%) | 0 (0.0%) | 1 (20.0%) |  |
|  | DUX4 p.I65N + ETV6_Q12del | 1 (9.1%) | 0 (0.0%) | 0 (0.0%) |  |
|  | ERG::LINC01423 | 0 (0.0%) | 1 (7.1%) | 0 (0.0%) |  |
|  | total | 1 (9.1%) | 2 (14%) | 1 (20.0%) |  |
| MEF2D | MEF2D::BCL9 + ATM p.P604S | 1 (9.1%) | 0 (0.0%) | 0 (0.0%) |  |
| TCF3::PBX1 | TCF3::PBX1 + TCF3_S338fs*10 | 0 (0.0%) | 1 (7.1%) | 0 (0.0%) |  |
| ZNF384 | EP300::ZNF384 | 0 (0.0%) | 1 (7.1%) | 0 (0.0%) |  |
| B-other | Unclassified | 1 (9.1%) | 0 (0%) | 1 (20.0%) |  |

<sup>1</sup>two patients had missing information on MRD (1 hyperdiploid and 1 Ph-like); <sup>2</sup>Final classification; <sup>3</sup>Kruskal-Wallis rank sum test; <sup>4</sup>Fisher's exact test (by molecular subtype); <sup>5</sup>Classified as DUX4/Hyperdiploid; <sup>6</sup>Novel fusion transcripts. In bold are depicted the high-risk alterations identified in patients initially classified as intermediate-risk according to the
protocol guidelines.

Finally, given the association between *CRLF2* rearrangements and Hispanic ancestry^29^, we assessed the prevalence of *P2RY8::CRLF2* in additional samples using RT-PCR. This fusion was detected in 4 of 56 patients (7%) and, as expected, was enriched in relapsed cases (50% vs 13%, Supplementary Material S2).

## Discussion

Acute lymphoblastic leukemia (ALL) is the most common pediatric cancer, and its treatment success increasingly depends on precise molecular characterization. Comprehensive profiling of chromosomal alterations, genetic mutations and gene expression patterns is essential for refining risk stratification and guiding precision and targeted therapies. However, the heterogeneity of ALL, coupled with the increasing complexity of molecular and bioinformatic analyses, presents significant challenges for achieving a complete and standardized molecular classification. Integrating multiple genomic and transcriptomic approaches is therefore critical to improve disease subtyping, identify novel alterations, and enhance precision medicine strategies. In this study, we conducted a comprehensive molecular characterization of childhood B-ALL in Argentine patients using whole-transcriptome sequencing (RNA-seq) on bone marrow aspirates at diagnosis. Our findings offer valuable insights into the molecular diversity of childhood B-ALL and support the incorporation of transcriptome sequencing into clinical diagnostics, prognosis assessment and precision treatment planning.

Identifying ALL genetic driver lesions and molecular subtypes at diagnosis is crucial for accurate risk stratification and treatment decision-making. However, its implementation, particularly in Latin American countries, remains challenging. The increasing number of ALL molecular subtypes and the diversity of genetic alterations demand large resources, including specialized personnel, infrastructure, funding and time, which are often limited in this region. For instance, the ALLIC-BFM-2009 protocol, which recruited 6,187 patients from 13 resource-limited countries, reported that over 60% of patients lacked cytogenetic and/or molecular results, leading to incomplete molecular characterization of the disease^19^. Furthermore, most molecular subtyping tools were primarily developed using cohorts from the U.S. and Western Europe, raising concerns about their classification accuracy and clinical relevance in populations with different genetic background and environmental exposures. Previous studies have also shown that therapeutic response rates are highly variable and tend to be lower in patients from low– and middle-income countries^5,6^.

Expanding molecular and genetic subtyping in underrepresented populations will require improved access to technical and financial resources. In this context, our study is the first to provide a detailed molecular profiling of Argentine patients using freely available bioinformatic tools. While the combination of multiple tools improved classification rates, we demonstrated that the analysis of a selected set of genes and fusion transcripts enabled effective identification of prognostic molecular alterations and subtypes without requiring advanced computing infrastructure and programming expertise, making them promising tools for implementation in clinical settings.

The study of fusion genes is critical for accurate ALL diagnosis, prognosis, and risk stratification. In this study we used STAR-Fusion, a tool that demands high computational resources, and RNAmut, which requires a pre-defined list of protein-coding genes, resulting in the need of fewer computational resources. We observed that both tools failed to detect all fusions, and overall, the agreement between them was low. By employing multiple fusion detection tools, we significantly increased the detection rate of chimeric transcripts. These findings underscore the need for a complementary, multi-tool strategy to ensure a more comprehensive molecular characterization of leukemic samples. Similarly, Vicente Garces *et al.* found that none of the five fusion-calling pipelines tested achieved perfect sensitivity and precision^30^. In resource-limited contexts, this approach is particularly valuable when integrated with user-friendly, open-access pipelines.

We also confirmed that the analysis of gene expression patterns from RNA-seq data can provide additional supporting evidence to validate the molecular subtype and the presence of fusion genes. For example, in our study, while STAR-Fusion detected the *MEF2D::BCL9* chimera, RNAmut did not. To resolve this discrepancy, we analyzed the expression of *HDAC9*, as it has been reported that cells and patients with the *MEF2D::BCL9* fusion have a markedly high expression of HDAC9^31,32^. As expected, we observed high *HDAC9* expression in this patient, supporting the classification as MEF2D subtype and the presence of *MEF2F::BCL9*. This highlights how integrating molecular characterization, subtyping and gene expression data can strengthen molecular diagnoses.

RNAmut is a user-friendly tool originally developed for acute myeloid leukemia. To our knowledge, our study is the first application of RNAmut to ALL samples. We constructed a custom gene index aimed at identifying clinically relevant SNVs, InDels and gene fusions in ALL. This allowed us to identify novel SNVs in genes previously reported as mutated in ALL or other hematological malignancies: *CREBBP*, *CSF3R* and *DUX4*. *In silico* modeling of the novel SNVs predicted the disruption of normal protein folding and/or activity. The SNV in CREBBP (p.G1542V) was found in one patient who was classified with an intermediate-risk disease who relapsed at 33 months; and was found in co-occurrence with a truncating oncogenic mutation in *NF1*. In agreement with this, *CREBBP* mutations have been previously associated with a high incidence of relapse in ALL patients^25^, whereas *NF1* mutations do not seem to change overall survival in ALL^33^.

The *CSF3R* p.G147R variant was identified in an intermediate-risk patient with a hyperdiploid subtype and MRD<0.1% at day 15, co-occurring with the novel *DBF4B::EFTUD2* fusion. Our *in silico* analysis suggested that the *CSF3R* mutation affects protein function. While *CSF3R* mutations are rare in B-ALL^34,35^, truncating and activating mutations in this gene are known drivers in chronic neutrophilic leukemia and atypical chronic myeloid leukemia^36^. In addition, mutations in the intracellular domain of CSF3R have been associated with specific therapeutic vulnerabilities, including sensitivity to dasatinib and resistance to ruxolitinib^37^, suggesting potential clinical relevance. The novel chimera *DBF4B::EFTUD2* results from an in-frame fusion of two protein-coding genes. DBF4B is a key regulator of CDC7 kinase, which is involved in DNA replication and response to replication stress. Although the biological function of the chimeric protein remains unknown, the patient exhibited near-null expression of wild-type DBF4B, potentially disrupting replication processes, as previously reported in DBF4-deficient cells^38^. The fusion partner EFTUD2 is a spliceosomal GTPase. Notably, we detected high expression of wild-type *EFTUD2* in this patient, consistent with its reported oncogenic role in hepatocellular carcinoma^39^, and suggesting that *EFTUD2* may also contribute to leukemogenesis in ALL.

The third novel SNV we identified was DUX4 p.I65N, located within one of the protein’s homeodomains. Based on our predictions, this variant may impair DUX4 transcriptional activity. Notably, the patient carrying this variant showed the highest expression of *DUX4* within our cohort, despite DUX4 being normally expressed only in embryos, testes and thymus^40^. Interestingly, previous studies have shown that overexpression of full-length or C-terminal-truncated DUX4 can be toxic to cells^41^. This apparent discrepancy may be explained by the predicted impaired DNA-binding affinity of the DUX4 p.I65N variant, which could allow cells to tolerate high levels of full-length *DUX4* without experiencing toxic effects. This variant co-occurred with ETV6_Q12del, a predicted spliced variant that was not detected by CTAT-Mutations. The functional relevance of this variant remains to be studied. The patient having these mutations was classified as intermediate-risk with MRD<0.1%, had a DUX4 subtype, and has neither relapsed nor died as of the latest analyses, suggesting a milder disease.

Finally, we identified a novel fusion involving a pseudogene/long non-coding RNA: *SCAF8::FER1L4*. This fusion is transcribed into a chimeric RNA of 6,907 nucleotides, comprising the N-terminal region of SCAF8 (714 amino acids), 11 amino acids from FER1L4, and a premature stop codon. *FER1L4* is annotated as a pseudogene in Ensembl (ENSG00000088340.11; last access: april 5th, 2025) but has been described as a long non-coding RNA by several studies. Initially identified in gastric cancer, *FER1L4* has since been reported to be an oncogene by modulating the PI3K/AKT pathway in various cancer cells, including osteosarcoma, hepatocellular carcinoma, and colon cancer. It plays critical roles in numerous processes such as cell growth, apoptosis, migration, and invasion^42^. The biological activity of this novel fusion in ALL and other malignancies remains to be studied.

Although our study was limited by the small number of relapses and a median follow-up of 32 months, we observed that all patients who relapsed harbored genetic alterations at diagnosis previously associated with high risk of relapse in other cohorts.

Altogether, RNA-seq analysis of leukemic bone marrow enabled molecular subtyping in 93.7% of B-ALL patients and facilitated the identification of both known and novel molecular alterations. These findings reinforce the value of transcriptome-based approaches as powerful tools for comprehensive molecular diagnostics, particularly in resource-constrained settings.

## Conclusions

This study presents the first comprehensive analysis of the molecular landscape in Argentine pediatric and adolescent patients diagnosed with B-ALL. The use of whole transcriptome sequencing (RNA-seq) on diagnostic bone marrow aspirates enabled the integrative assessment of multiple molecular alterations, including gene expression profiles, small sequence variations, and gene fusions. Importantly, the use of a multi-tool strategy combined with user-friendly and open-access pipelines represents a significant advancement for implementing comprehensive molecular diagnostics in resource-limited countries. These findings add significant clinical value, offering potential improvements in risk stratification and the identification of therapeutic targets, particularly in cases where traditional methods fail to define a molecular subtype. Further studies are warranted to evaluate whether the detection of these alterations could aid in the early detection of relapses, precision medicine or in selecting candidates for clinical trials.

## Data availability

Transcriptome sequencing data is available at the European Nucleotide Archive repository under the Accession PRJEB80172. Variants, fusion transcripts, molecular subtypes, gene lists and primers are listed in the file “Supplementary Material S1”. Additional methods are detailed in the Supplementary Material S2.

## Author contributions

Conceptualization, Maria Sol Ruiz and Javier Cotignola; Formal analysis, Maria Sol Ruiz and Javier Cotignola; Funding acquisition, Geraldine Gueron and Javier Cotignola; Investigation, Maria Sol Ruiz, Daniel Avendaño, Ignacio Gomez Mercado and Maria Laura Lacreu; Methodology, Maria Sol Ruiz and Ezequiel Sosa; Resources, Maria Cecilia Riccheri, Virginia Schuttenberg and Luis Aversa; Software, Maria Sol Ruiz and Ezequiel Sosa; Supervision, Elba Vazquez, Geraldine Gueron and Javier Cotignola; Writing – original draft, Maria Sol Ruiz and Javier Cotignola; Writing – review & editing, Maria Sol Ruiz and Javier Cotignola.

## Acknowledgements

We thank members of the GATLA (Grupo Argentino de Tratamiento de Leucemias Agudas) working group, and all physicians, biochemists and patients involved in this study. We thank Santiago Di Lella, PhD., for his contributions in the structural analysis of novel variants and visualization of protein structures. We thank Maria Mercedes Abbatte for her contributions in the processing of samples for transcriptome sequencing.

## Ethics approval statement

All procedures involving human participants complied with the ethical guidelines outlined by the Institutional Review Boards (IRB) and with the Declaration of Helsinki. Additionally, the study protocol received approval from Argentina’s National Drug, Food and Technology Administration (ANMAT).

